# Autostent: A Semi-Automated Approach to Designing Customized 3D-Printed Oral Radiation Stents for Patients with Head and Neck Cancer

**DOI:** 10.1101/2023.09.26.23296170

**Authors:** Anshuman Agrawal, Mohamed M. Zaid, Millicent A. Roach, Lianchun Xiao, Mark S. Chambers, Eugene J. Koay

## Abstract

Oral stents may reduce toxicities during radiation therapy for head and neck cancer (HNC). Although customized 3D-printed oral stents are more quickly fabricated and non-inferior (in terms of patient reported outcomes) compared to conventionally-fabricated stents, the design process is still relatively time-consuming, unstandardized, and requires experienced technicians. We hypothesized that semi-automating the 3D-printed stent design process can reduce design time and standardize the workflow. Using oral stent design principles established over decades by oral oncologists, we developed a customized computer program (Autostent) using MATLAB to semi-automate the design process. We then compared a previously described method utilizing non-automated computer-aided design with Autostent. Three users designed stents for four patients with HNC enrolled on a prospective observational study. These patients were selected based on differing dental anatomies, and each user designed stents for each patient thrice, using both the non- and semi-automated methods. The design time and stent volumes for the two methods were statistically analyzed. Semi-automation was found to significantly reduce the average design time by 23.6 minutes (51.2%, p=0.001), independent of user, dental anatomy, and trial number. Additionally, semi-automation reduced the average stent volume by 4.33 mL (12.9%, p=0.016, univariate analysis). While this was not statistically significant after accounting for the other experimental variables (p=0.40, multivariate analysis), semi-automation did reduce the variability in the stent volume across users (overall standard error of the mean reduced by 40%). Thus, the semi-automated workflow significantly reduced the design time and the variability in the stent volume across users compared to the non-automated workflow. This may lead to potential cost benefits, standardization of the device, and increased population-wide access to a device that could help reduce toxicities for HNC.

## Introduction

Radiation therapy is an effective, non-invasive technique shown to improve overall survival and local control in patients with head and neck cancer (HNC), the 8th most common cancer globally.^1-3^ However, the delivery of higher radiation doses is counterbalanced by toxicities to the healthy tissue, notably radiation-induced oral mucositis.^4-6^ These toxicities can greatly reduce patients’ quality of life and lead to treatment breaks.^6-8^ Toxicity sparing devices such as oral stents have been utilized for decades to improve the therapeutic index of radiotherapy.^9-14^ The conventional approach to manufacture fully-customized oral stents requires multiple in-person patient visits and a team of radiation oncologists and oral-maxillofacial surgeons with oncology specific training to handcraft these stents.^15^ This increases the cost and paradoxically limits scalability of these devices in high-volume treatment centers, while leaving low-volume centers without an option for fully-customized stents altogether.

However, by using a previously-described workflow to design customized, 3D-printed, mouth-opening-tongue-depressing (MOTD) oral stents, the fabrication time can be reduced from approximately 48 hours to 8 hours.^15-17^ Moreover, the resulting 3D-printed stents are non-inferior to conventionally-fabricated stents in terms of patient-reported outcomes.^15^ This workflow involves the utilization of an intraoral 3D scanner at the point-of-care to scan the patient’s dental anatomy, Computer-Aided Design (CAD) software to manually design the stent, and a 3D printer to print the customized stent. Even with the improved turnaround time compared to the conventional approach (48hrs vs. 8hrs), the stent design process is still relatively time-consuming and requires extensive experience with the CAD software.^15^ Moreover, the quality standardization is challenging, as design decisions are primarily based on the designer’s experience. Therefore, to address this unmet need for a faster and more standardized design approach, we describe a semi-automated approach using a customized software (Autostent). We hypothesize that using such an approach can reduce the design time, standardize the workflow, and increase consistency in the stent volume, when compared to the original, non-automated method. This could lead to increased access to fully-customized oral stents in radiation treatment centers, globally.

## Materials and Methods

### I. Semi-Automation

The design workflow was semi-automated using MATLAB R2020a (Mathworks Inc.). MATLAB’s Application Designer was used to create the user interface for Autostent (*Figure 1a*) and the workflow was standardized into 9 steps (*Figure 1b* visualizes each step’s output).

**Figure 1.**
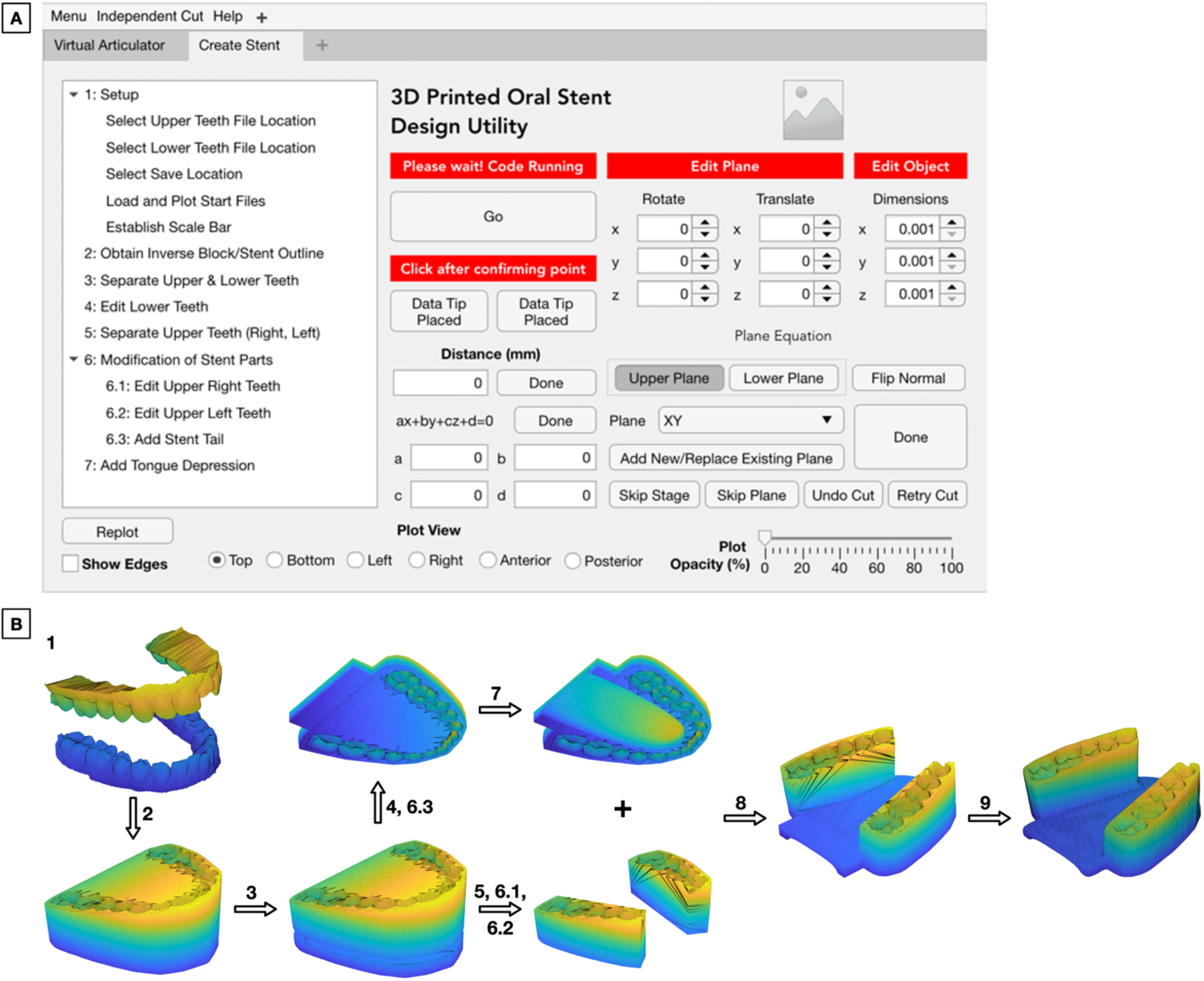
Autostent’s user interface (A) guides users through the workflow in a standardized, step-wise manner. Outputs of each step of the semi-automated workflow (B). Steps 1-7 are completed on Autostent (listed on the left in A). Steps 8-9 are completed on Autodesk MeshMixer.

#### Step 1

The patient’s 3D maxillary and mandibular intraoral scans are imported and the 20mm inter-incisal opening is used to define a scale bar.

#### Step 2

Best-fit planes to the maxilla and mandible are generated using Principal Component Analysis. The maxillary plane is positioned at 60% of the height (from the upper gums), whereas the mandibular plane is positioned at 40% of the height (from the lower gums). The user can fine-tune these planes. The combined convex hull of the patient’s maxillary and mandibular structures is computed, expanded outwards by 1.5%, and sliced off at the defined planes. The patient’s teeth are subtracted from the sliced convex hull to create the customized dental impression. The slicing of the 3D mesh is achieved using Constructive Solid Geometry functions in the iso2mesh toolbox.^18^ Each plane cut is implemented as a Boolean subtraction between the mesh and a cube (one face angled to correspond to the plane).

#### Step 3

The upper and lower teeth are separated using a plane initially positioned 2mm above the lower central incisor (identified in step 1 while defining the scale bar) and whose slope is based on the mandibular plane defined in step 2.

#### Step 4

The third molars are removed from the lower stent.

#### Steps 5, 6.1, 6.2

The left and right upper teeth supports are created by modifying the upper stent using 3 pre-defined, user-customizable vertical planes for each side. Although unneeded, the supports can be further modified in steps 6.1-6.2.

#### Step 6.3

The stent tail required to depress the tongue is added by merging the lower stent with a user-customizable box extending 10mm beyond the second molar.

#### Step 7

A standard tongue-like mesh is manually positioned and subtracted from the lower stent to create space for the patient’s tongue.

After each step, Autostent saves the resulting 3D mesh as an STL (Stereolithography) file to a user-defined folder to prevent data loss. These are used to complete steps 8-9 in Autodesk Meshmixer.

#### Step 8

The left and right upper stent supports and the lower stent are merged.

#### Step 9

Sharp edges are smoothed and the file is exported for 3D printing.

### II. Experimental Design

#### Patient Population

We evaluated four patients with unique dental anatomies (Class III Malocclusion, Edentulous, Hypodontic, and Normal Occlusion) who received definitive radiotherapy for primary HNC between 2018-2019 and were referred to the oral oncologist for fabrication of standard oral stents. The study was conducted under Institutional Review Board approved protocol (2017-0269).

#### Stent Design

The patients’ maxillary and mandibular anatomy was scanned as STL files using an intraoral scanner (Trios 3shape) from their articulated stone models (used to originally fabricate the standard stents). Three independent researchers (Users 1 (AA), 2 (MR), 3 (MZ)) designed three stents for each of the four patients using: 1) the original, non-automated design method utilizing Autodesk Meshmixer, and 2) the semi-automated method utilizing Autostent. The design time was recorded and the stent volume was measured.

#### Statistical Analysis

Linear mixed models, including patient dental anatomy as random effects, were fitted to evaluate whether the differences in time and in volume between the two methods (per user per trial for each patient) were statistically significant (p<0.05). To adjust for the effects of users and trials, this was repeated after adding users and trials as covariates (multivariate analysis). ANOVA analysis and two-sample t-tests were used to compare the volumes between the two methods for each user.

## Results

### I. Design Time

The analysis indicated that designing stents was significantly faster using Autostent (mean difference = 23.6 min, 95% CI [17.6, 29.7], p=0.0016, *Figure 2a*). After adjusting for the effects of users and trials, the difference was still statistically significant (p=0.0009).

**Figure 2.**
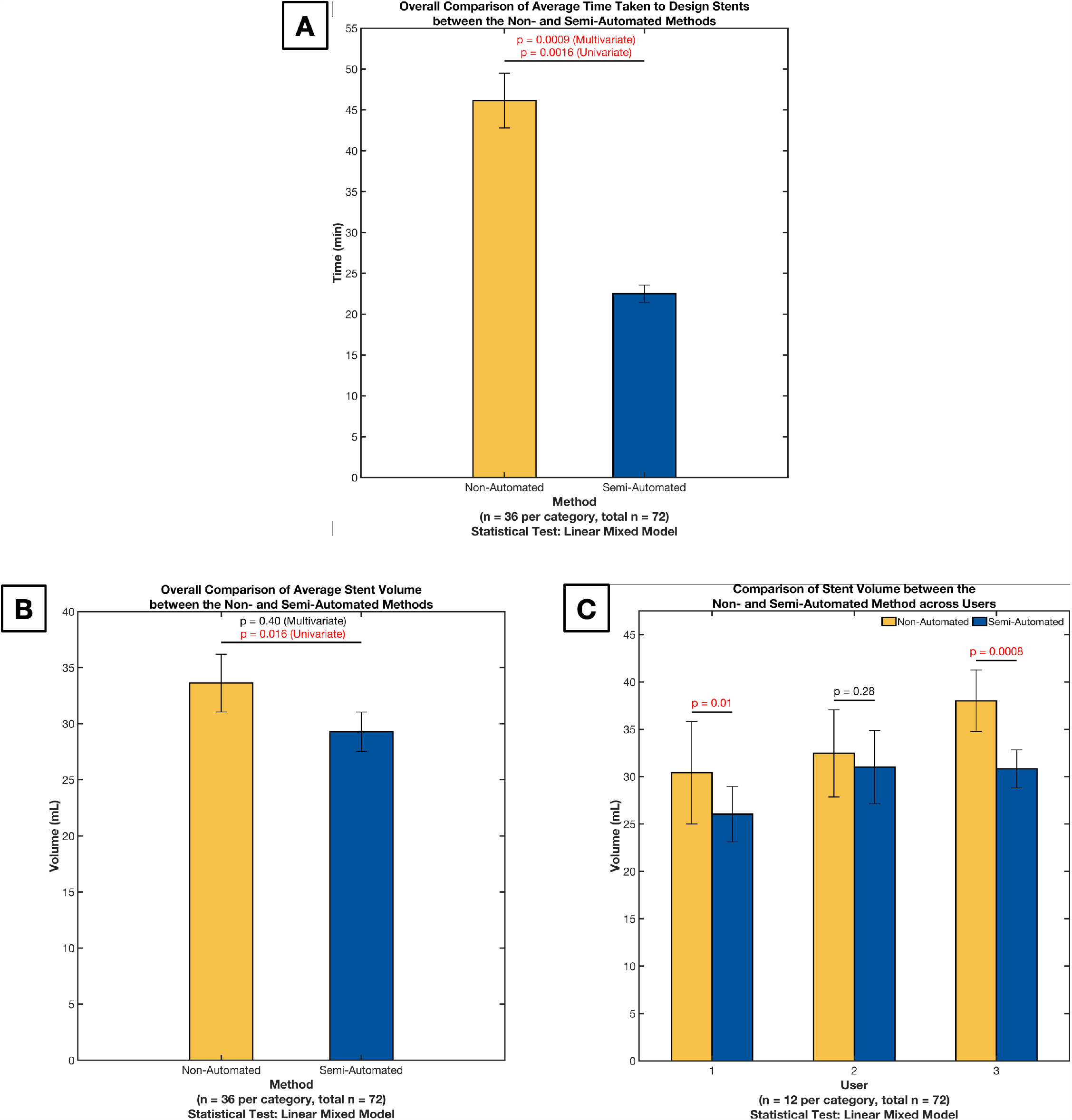
Results comparing the Non- and Semi-Automated methods using the design time (A), stent volume (B), and stent volume across users (C). The error bars represent the 95% confidence interval for the mean.

### II. Stent Volume

The univariate analysis indicated that stents designed using Autostent had significantly lower volumes (mean difference = 4.33 mL, 95% CI [2.20, 6.47], p=0.016, *Figure 2b*, visualized in *Figure 3*). After adjusting for the effects of users (p<0.0001) and trials (p=0.75), the difference was no longer statistically significant (p=0.40). Investigating the stent volumes between the two methods for each user (*Figure 2c*) indicated that semi-automation resulted in lower variability for all users (standard error of the mean was reduced by 1.1, 0.3, and 0.6 for users 1, 2, and 3, respectively, and by 0.4 overall).

**Figure 3.**
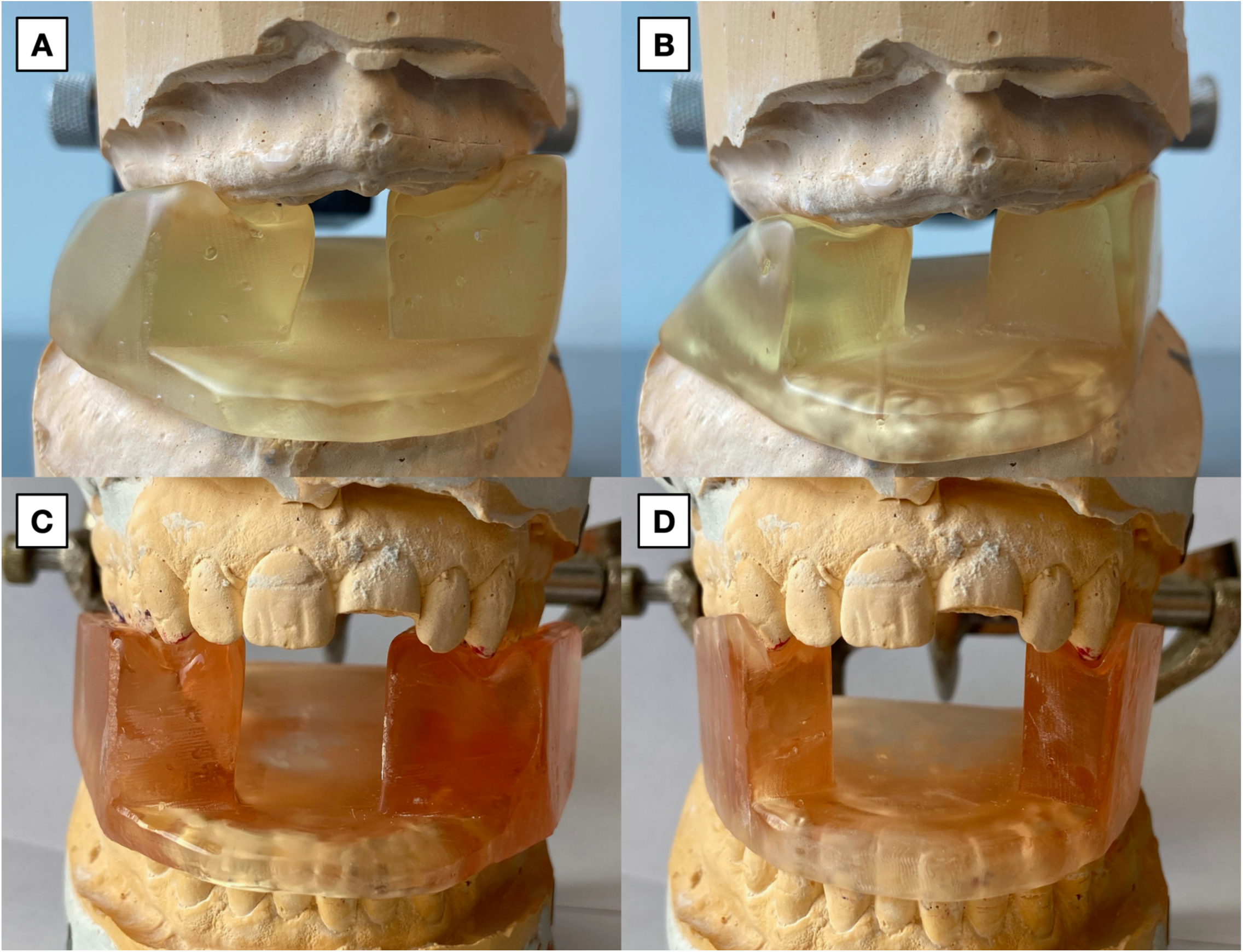
3D printed stents designed using the non-automated (A, C) and semi-automated (B, D) method for the edentuluous (A-B) and normal occlusion (C-D) dental anatomies. The boundary of the stent designed using the semi-automated method is more compact, thus reducing the volume and potentially resulting in a more comfortable fit.

## Discussion

The clinical conundrum of treating HNC patients with high-dose radiation while trying to minimize radiation-induced toxicities may be partially addressed using customized oral radiation stents that can help improve the therapeutic index.^9-14^ However, designing these stents is time-consuming and costly, with the design being unstandardized and designer-specific. The proposed semi-automated approach targets these avenues for improvement and, compared to the manual approach, reduces the average design time by 23.6 minutes (51.2%) and the variability in the stent volume across users by 40% (standard error of the mean was reduced by 0.4 overall).

The study was limited by the sample size of 4 unique patient cases and 3 users. This likely led to insufficient power in the stent volume analysis. Incorporating a wider variety of dental anatomies and users of varied skill levels might have led to more representative results. Future work will focus on incorporating machine learning algorithms to automatically identify dental landmarks and fully automate the design process, and on removing the dependence on existing CAD software by completing all the steps within Autostent.

In conclusion, the semi-automated workflow significantly reduces the time taken to design an MOTD oral stent (for four common dental anatomies). Moreover, in addition to potentially reducing the cost associated with fully-customized stents, Autostent also standardizes the design workflow and increases the consistency in the stent volume among different users. Especially given the increased usage of oral stents by patients undergoing radiation therapy for HNC, from 36% in 1992 to 67% in 2013, these advances represent a significant step towards enabling large-scale clinical implementation and providing population-wide access to fully-customized oral stents.^19^

## Data Availability

Research data are not available at this time.

